# Measuring odds of various COVID-19 infection prevention & control measures among the contacts traced during trace test and quarantine activities at district Quetta (An un-matched case control study)

**DOI:** 10.1101/2021.07.25.21261084

**Authors:** Muhammad Arif, Ehsan Larik, Abid Saeed, Muhammad Abdullah

## Abstract

**Background:** The second wave of COVID-19 pandemic has started globally, right now globally 220 countries are infected and a total of 71351695 confirmed cases and 1612372 deaths due to COVID-19 has been reported so far^1^. As of today i.e. (16^th^ Dec 2020) a total of 11430955 new COVID-19 confirmed cases have been reported across the south-east asia^1^. These cases are showing an increasing trend in all the Asian countries including Pakistan^1^. Across Pakistan till date 440787 new confirmed COVID-19 cases have been reported across the Pakistan showing a doubling time of 10.63 days (95% C.I 9.68-11.8), while a total of 8832 new deaths have been reported across the Pakistan making the double time for death as 11.11 days (95% C.I 4.04-14.86) ^1^. Till the development and availability of a vaccine the only tools that can help prevent the spread of COVID-19 are IPC measures violating them can result in a quick spread across the population^3^.

This study was conducted to assess the odds of various COVID-19 IPC measures among the *<u>Contacts</u>* of an index COVID-19 case traced by Provincial Disease Surveillance & Response unit Quetta.

**Methodology:** *Sample Size & Sampling technique:* Using the detailed epidemiological reports of 600 COVID-19 **<u>*contacts*</u>** identified during the trace test and quarantine field activities form 1^st^ October till 30^th^ October 2020 in district Quetta, from this data a sample of **300** individuals was selected for this study using ***Simple*** random sampling technique.

*Study Design:* Considering different exposure rates and pandemic situation an ***Un-matched Case control study*** study was conducted where **Cases** were defined as “Every PCR positive contact (*Symptomatic or asymptomatic*) for any index case” similarly **Controls** were defined as “Every PCR negative contact (*Symptomatic or asymptomatic*) for any index case who was home quarantined for 14 days based on suspicion by PDSRU team. A set ratio of 1:2 for cases & controls respectively was used for this study.

*Results:* The odds for various IPC measures like *Knowingly and intentionally Contacted with a COVID-19 positive case, Family member of the index COVID-19 case, Knowingly and intentionally received an object handed over by a COVID-19 Positive case, Touched the same surface/surfaces after it was touched by the index case, Not doing regular Hand washing, Knowingly and intentionally did not follow the government SOPs of Social Distancing During encounter with a positive symptomatic case, Knowingly and intentionallydid not Follow the government SOPs of Social Distancing During sharing of bedroom and toilet with positive symptomatic case, used the same vehicle after it was used by the COVID-19 index case, Spoke with Positive COVID-19 index case for more than 15mins few days before catching the disease, Individual did not use a face mask during all of his contact episodes with the positive index case, Participating in gathering or social events* were all found to be poorly followed by the PCR positive contacts.

## Introduction

The second wave of COVID-19 pandemic has started globally, right now globally 220 countries are infected and a total of 71351695 confirmed cases and 1612372 deaths due to COVID-19 has been reported so far^1^. As of today i.e. (16^th^ Dec 2020) a total of 11430955 new COVID-19 confirmed cases have been reported across the south-east asia^1^. These cases are showing an increasing trend in all the Asian countries including Pakistan^1^. Across Pakistan till date 440787 new confirmed COVID-19 cases have been reported across the Pakistan showing a doubling time of 10.63 days (95% C.I 9.68-11.8), while a total of 8832 new deaths have been reported across the Pakistan making the double time for death as 11.11 days (95% C.I 4.04-14.86) ^1^. Till the development and availability of a vaccine the only tools that can help prevent the spread of COVID-19 are IPC measures violating them can result in a quick spread across the population^3^.

Provincial Disease Surveillance & response unit (PDSRU) has always been a first responders to each out break across Balochistan province and currently it is a focal point for trace test and quarantine (TTQ) strategy across Balochistan. PDSRU is run by trained Field epidemiologists trained by Field Epidemiology and laboratory Training program Pakistan. So as ever PDSRU responded to the COVID-19 second wave and took active part in implementing the trace test and Quarantine (TTQ) activities across Balochistan province including District Quetta and has recorded detailed epidemiological reports for every case in the field.

### Literature review

Infection prevention and control (IPC)measures for COVID-19 like keeping 6 feet social distancing, wearing face mask, avoiding gathering and regularly washing hands were all proved vitals in decreasing the transmission rates among the communities ^4,8,10,11^.In another study done by Hsiang S and others observed that the ongoing anti-contagion policies have already substantially reduced the number of COVID-19 infections observed in the world today according to their calculations all policies of SOPs and IPC measured when properly implemented slowed the average growth rate of infections by − 0.252 per day (SE= 0.045, 164 p< 0.001) in China, − 0.248 (SE= 0.089, p< 0.01) in South Korea, − 0.24 (SE= 0.068, p< 0.001) in 165 Italy, − 0.355 (SE= 0.063, p< 0.001) in Iran, − 0.123 (SE= 0.019, p< 0.001) in France and − 0.084 166 (SE= 0.03, p< 0.01) in the US^2^.

In another study done by Lai S and the others where they predicted the infection rates and quantified the impact of various non-pharmacological interventions (NPI) according their calculations Without NPIs, their model predicted the number of cases of COVID-19 to increase rapidly across China, with a 51-fold (IQR 33–71) increase in Wuhan, a 92-fold (58–133) increase in other cities in Hubei province and a 125-fold (77–180) increase in other provinces by 29 February 2020. However, the apparent effectiveness of different interventions varied. Nevertheless, if intercity travel restrictions had been implemented, cities and provinces outside of Wuhan would have not received more cases from Wuhan, and the affected geographical range would not have expanded to the remote western areas of China. In general, they estimated that the early detection and isolation of cases quickly and substantially adopting IPC measure more infections were controlled like contact reduction and social distancing measures across the country. However, without the contact reduction intervention, in the longer term the epidemics would have increased exponentially across regions. Therefore, collective NPIs would bring about the strongest and most rapid effect on containment of the COVID-19 outbreak, with an interval of about one week between the introduction of NPIs and the peak of the epidemic^3^.

In another study by Flaxman S and the others showed that major non-pharmaceutical interventions-and lockdowns in particular-have had a large effect on reducing transmission in Europeans. Continued intervention should be considered to keep transmission of SARS-CoV-2 under control^4^.

The waves of COVID-19 will continue to repeat, like wise every sector, profession and every human conduct will always remain prone to it till the production and availability of vaccine to the common people and almost 60% herd immunity is achieved^11, 12^. In these crisis situations trends towards new normal life must be focused and IPC measures should be made part of routine^5, 6,7,14,15^.

### Operational Definition of “Contact”

Contact was defined as a person who have had contact, without effective protection regardless of duration of exposure, with 1 or more persons with suspected or confirmed COVID-19 any time starting 2 days before onset of symptoms in persons with a suspected or confirmed case, or 2 days before sampling for laboratory testing of asymptomatic infected persons^10^.

### Problem statement

COVID-19 is a highly contagious disease till the development and availability of COVID-19 vaccine the waves of this pandemic will continue to occur repeatedly hence each wave could potentially reach to new heights of infectivity and mortality.

### Rationale

So far no published literature has studied the odds for COVID-19 infection prevention and control (IPC) measures among the common masses of developing countries like Pakistan where literacy rates are low poverty and population is high as a result huge number of family members shares a single room for living.

### Objectives

- To assess the odds for certain anti-COVID-19 IPC measures among the *<u>Contacts</u>* of an index COVID-19 case traced by Provincial Disease Surveillance & Response unit Quetta.
- To provide evidence based recommendations for risk communication to the local context via media cell of the Health department of Balochistan so that certain COVID-19 IPC measures are adopted and focused by every resident of Balochistan in their daily life activities till the availability of anti-COVID-19 vaccine.

### Research Question

Q: How are the odds for certain anti-COVID 19 IPC measures among the contacts of an index case identified during test, trace and quarantine (TTQ) activities?

Q: Which anti-COVID-19 IPC measures needs more focus in order to decrease the transmission of COVID-19 among the residents of Quetta?

### Hypothesis

#### H_0_

Both COVID-19 PCR Positive and PCR negative contacts of an index case practice certain anti COVID-19 IPC measures equally in their daily life like *Knowingly and intentionally Contacted with a COVID-19 positive case, Family member of the index COVID-19 case, knowingly and intentionally received an object handed over by a COVID-19 Positive case, Touched the same surface/surfaces after it was touched by the index case, Not doing regular Hand washing, Knowingly and intentionally did not follow the government SOPs of Social Distancing During encounter with a positive symptomatic case, Knowingly and intentionally did not follow the government SOPs of Social Distancing During sharing of bedroom and toilet with positive symptomatic case, used the same vehicle after it was used by the COVID-19 index case, Spoke with Positive COVID-19 index case for more than 15mins few days before catching the disease, Individual did not use a face mask during all of his contact episodes with the positive index case, Participating in gathering or social events*.

#### H_a_

Both COVID-19 PCR Positive and PCR negative contacts of an index case ***<u>do not</u>*** practice certain anti COVID-19 IPC measures equally in their daily life like *Knowingly and intentionally Contacted with a COVID-19 positive case, Family member of the index COVID-19 case, Knowingly and intentionally received an object handed over by a COVID-19 Positive case, Touched the same surface/surfaces after it was touched by the index case, Not doing regular Hand washing, Knowingly and intentionally did not follow the government SOPs of Social Distancing During encounter with a positive symptomatic case, Knowingly and intentionallydid not Follow the government SOPs of Social Distancing During sharing of bedroom and toilet with positive symptomatic case, used the same vehicle after it was used by the COVID-19 index case, Spoke with Positive COVID-19 index case for more than 15mins few days before catching the disease, Individual did not use a face mask during all of his contact episodes with the positive index case, Participating in gathering or social events*.

## Methodology

### Sample Size & Sampling technique

PDSRU Quetta’s Field epidemiologist team recorded detailed epidemiological reports of 600 COVID-19 ***<u>contacts</u>*** from during the trace test and quarantine field activities form 1^st^ October till 30^th^ October 2020 in district Quetta, from this data a sample of **300** individuals was selected for this study using ***Simple random sampling technique***. Using the following formula of sample size where C.I of 95%, 0.5 Population proportion(p), 0.04 Margin of error (e), 600 Population size of the total contacts traced during the field activities of TTQ by PDSRU Field epidemiologist team from 1 ^st^ October till 30^th^ October 2020, 0.025 alpha divided by 2 and a z-score of 1.96 values were used for the calculation of sample size of 300.

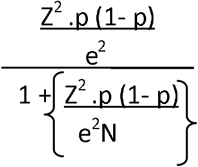

### Study Design

Considering different exposure rates and pandemic situation an ***Un-matched Case control study*** study was conducted where **Cases** were defined as “Every PCR positive contact (*Symptomatic or asymptomatic*) for any index case” similarly **Controls** were defined as “Every PCR negative contact (*Symptomatic or asymptomatic*) for any index case who was home quarantined for 14 days based on suspicion by PDSRU team. A set ratio of 1:2 for cases & controls respectively was used for this study.

### Data Collection Tool

An interview using structured questioner was conducted with every individual during data collection.

### Analysis Plan

Epi-info software was used; descriptive statistics for age, sex, educations status, Blood groups, Co-morbidities, BCG & Seasonal flue vaccination status of the study participants were summarized using frequency tables, while 2×2 contingency table was used for the calculation Odds ratios.

## Results

### Descriptive statistics

#### Socio-demographic Characteristics of Respondents

Total 300 contacts were included in this study and their age distribution was as 195 respondents (65%) were in age category ranging from 1-35 years, 101 (34%) were in range of >35 years. Similarly 100 (33%) of the study participants were found to be COVID-19 PCR positive and were considered as cases while 200 (67%) were found to be COVID-19 PCR negative and were taken as controls.

Gender distribution of the participants showed that 197 (66%) individuals were male while 103 (34%) were females.

For Ethnicity of respondents, 125 (42%) were Pashtons, 95 (32%) were Baloch, 55(18.3 %) were Brahvi, 15 (05%) were Punjabi and 10 (3.3%) respondent were from Hazara ethnic group. Some of the other socio demographic characteristics of the participants are summarized as below:

**Table No:** 01-Socio Demographic Characteristics of the study Participants:

| S.NO | Socio-demographic Characteristics of the study participants | Sub-Set | Percentages (%) | Number (n) | COVID-19 PCR +ve Contacts visited by PDSRU team (Cases) | COVID-19 PCR -ve Contacts visited by PDSRU team (Cases) |
| --- | --- | --- | --- | --- | --- | --- |
| 1 | Qualification | Un Educated | 49% | 39 | 13 | 26 |
|  |  | Primary | 9% | 45 | 15 | 30 |
|  |  | Metric | 20% | 27 | 9 | 18 |
|  |  | Intermediate | 17% | 120 | 40 | 80 |
|  |  | Graduation | 3% | 39 | 13 | 26 |
|  |  | Masters | 2% | 30 | 10 | 20 |
|  |  | Phd | 0% | 0 | 0 | 0 |
| 2 | Co-Morbidities | No Comorbidity | 90% | 270 | 90 | 180 |
|  |  | DM | 02% | 6 | 2 | 4 |
|  |  | Asthma | 04% | 9 | 3 | 6 |
|  |  | HTN | 04% | 12 | 4 | 8 |
|  |  | Hepatitis | 02% | 6 | 2 | 4 |
|  |  | Ovarian CA | 00% | 0 | 0 | 0 |
| 3 | Occupation | Health Care Providers | 20% | 60 | 20 | 40 |
|  |  | Others | 80% | 240 | 80 | 160 |
| 4 | BCG Vaccination status | BCG Vaccination NO | 63% | 189 | 63 | 126 |
|  |  | BCG Vaccination Yes | 37% | 111 | 37 | 74 |
| 5 | Seasonal flu Vaccination status | Seasonal Flu vaccination Yes | 7% | 21 | 7 | 14 |
|  |  | Seasonal Flu vaccination NO | 93% | 279 | 93 | 186 |
| 6 | Blood Groups of the study participants | A+ | 7.00% | 21 | 7 | 14 |
|  |  | A- | 4.00% | 12 | 4 | 8 |
|  |  | B+ | 27.00% | 81 | 27 | 54 |
|  |  | B- | 3.00% | 9 | 3 | 6 |
|  |  | AB+ | 2.00% | 6 | 2 | 4 |
|  |  | AB- | 1.00% | 3 | 1 | 2 |
|  |  | O+ | 1.00% | 3 | 1 | 2 |
|  |  | O- | 1.00% | 3 | 1 | 2 |
|  |  | unkown | 54.00% | 162 | 54 | 108 |

#### Inferential statistics

The following table summarizes the major findings of this study which are almost in line with the set hypothesis; this study is clearly showing that the odds of various COVID-19 infection prevention and control (IPC) measures studied among COVID-19 PCR positive contacts (cases) and COVID-19 PCR negative contacts (controls) were found to be significant as shown below:

**Table No:** 02-Odds ratios for the various Infection prevention and control measures studied in this study.

| **COVID-19 IPC Measures studied in this study** | **Responses** | **COVID-19 PCR Positive contacts (Case= 100)** | **COVID-19 PCR Negative contacts (Controls= 200)** | **O.R.** | **95% C.I** | **P-value** |
| --- | --- | --- | --- | --- | --- | --- |
| Knowingly and intentionally Contacted with a COVID-19 positive case. | Yes | 80 | 45 | 13.7 | 7.62-24.90 | 0.00 |
|  | NO | 20 | 155 |  |  |  |
| Family member of the index COVID-19 case | Yes | 25 | 9 | 7.07 | 3.25-15.86 | 0.00 |
|  | NO | 75 | 191 |  |  |  |
| Knowingly and intentionally received an object handed over by a COVID-19 Positive case. | Yes | 19 | 7 | 6.46 | 2.61-15.98 | 0.00 |
|  | NO | 81 | 193 |  |  |  |
| Touched the same surface/surfaces after it was touched by the index case | Yes | 70 | 55 | 6.15 | 3.62-10.43 | 0.00 |
|  | NO | 30 | 145 |  |  |  |
| Not doing regular Hand washing | Yes | 83 | 36 | 22.24 | 11.79-41.94 | 0.00 |
|  | NO | 17 | 164 |  |  |  |
| Knowingly and intentionally did not follow the government SOPs of Social Distancing During encounter with a positive symptomatic case. | Yes | 13 | 5 | 5.58 | 2.01-16.85 | 0.00 |
|  | NO | 87 | 195 |  |  |  |
| Knowingly and intentionally did not follow the government SOPs of Social Distancing During sharing of bedroom and toilet with positive symptomatic case. | Yes | 23 | 10 | 5.67 | 2.58-12.48 | 0.00 |
|  | NO | 77 | 190 |  |  |  |
| used the same vehicle after it was used by the COVID-19 index case | Yes | 63 | 16 | 19.58 | 10.19-37.60 | 0.00 |
|  | NO | 37 | 184 |  |  |  |
| Spoke with Positive COVID-19 index case for more than 15mins few days before catching the disease | Yes | 69 | 25 | 15.58 | 8.58-28.27 | 0.00 |
|  | NO | 31 | 175 |  |  |  |
| Individual did not use a face mask during all of his contact episodes with the positive index case | Yes | 91 | 69 | 19.19 | 9.11-40.41 | 0.00 |
|  | No | 9 | 131 |  |  |  |
| Participating in gathering or social events | Yes | 73 | 19 | 25.75 | 13.48-49.17 | 0.00 |
|  | No | 27 | 181 |  |  |  |

As shown in the above table the odds for *Knowingly and intentionally Contacted with a COVID-19 positive case* was 13.7 times more among the PCR Positive Contacts as compare to PCR negative Contacts (*p*=0.00,*C*.*I*=7.62-24.90), Similarly the odds of *being a Family member of the index COVID-19 case* was 7.07 times more among the PCR Positive Contacts as compare to the PCR negative Contacts (*p*=0.00,*C*.*I*=3.25-15.86), also the odds for *Knowingly and intentionally received an object handed over by a COVID-19 Positive case* was 6.64 times more among the PCR Positive Contacts as compare to PCR negative Contacts(*p*=0.00,*C*.*I*=2.61-15.98), where as the odds for *Touched the same surface/surfaces after it was touched by the index case* was 6.15 times more among the PCR Positive Contact as compare to PCR negative Contacts (*p*=0.00,*C*.*I*=3.62-10.43), more over the odds for *Not doing regular Hand washing* was 22.24 times more among the PCR Positive Contacts as compare to the PCR negative Contacts (*p*=00.00,*C*.*I*=11.79-41.94), while the odds for *Knowingly and intentionally did not follow the government SOPs of Social Distancing During encounter with a positive symptomatic case* was 5.58 times more among the PCR Positive Contacts as compare to the PCR negative Contacts (*p*=00.00,*C*.*I*=2.01-16.85),*also the odds for knowingly and intentionally did not Follow the government SOPs of Social Distancing During sharing of bedroom and toilet with positive symptomatic case* was 5.67 times more among the PCR Positive Contacts as compare to PCR negative Contacts(*p*=00.00,*C*.*I*=2.58-12.48), more over the odds for *used the same vehicle after it was used by the COVID-19 index case* was 19.58 times more among the PCR Positive Contacts as compare to PCR negative Contacts (*p*=00.00,*C*.*I*=10.19-37.60), also *the odds for Spoke with Positive COVID-19 index case for more than 15mins few days before catching the disease* was 15.58 times more among the PCR Positive Contacts as compare to the PCR negative Contacts (*p*=00.00,*C*.*I*=8.58-28.27), similarly the *odds for Individual did not use a face mask during all of his contact episodes with the positive index case* was 19.19 times more among the PCR Positive Contacts as compare to PCR negative Contacts(*p*=00.00,*C*.*I*=9.11-40.41), lastly the odds for *Participating in a gathering or social events* were 25.75 times more among the PCR Positive Contacts as compare to PCR negative Contacts(*p*=00.00,*C*.*I*=13.48-49.17).

## Discussion

The major findings of this study are almost in line with the set hypothesis, this study is clearly showing that the odds of various COVID-19 infection prevention and control (IPC) measures studied among COVID-19 PCR positive contacts (cases) and COVID-19 PCR negative contacts (controls) were found to be significant likewise the odds for *Knowingly and intentionally Contacted with a COVID-19 positive case* was 13.7 times more among the PCR Positive Contacts as compare to PCR negative Contacts (*p*=0.00,*C*.*I*=7.62-24.90), Similarly the odds of *being a Family member of the index COVID-19 case* was 7.07 times more among the PCR Positive Contacts as compare to the PCR negative Contacts (*p*=0.00,*C*.*I*=3.25-15.86), also the odds for *Knowingly and intentionally received an object handed over by a COVID-19 Positive case* was 6.64 times more among the PCR Positive Contacts as compare to PCR negative Contacts(*p*=0.00,*C*.*I*=2.61-15.98), where as the odds for Touched the *same surface/surfaces after it was touched by the index case* was 6.15 times more among the PCR Positive Contact as compare to PCR negative Contacts (*p*=0.00,*C*.*I*=3.62-10.43), more over the odds for *Not doing regular Hand washing* was 22.24 times more among the PCR Positive Contacts as compare to the PCR negative Contacts (*p*=00.00,*C*.*I*=11.79-41.94), while the odds for *Knowingly and intentionally did not follow the government SOPs of Social Distancing During encounter with a positive symptomatic case* was 5.58 times more among the PCR Positive Contacts as compare to the PCR negative Contacts (*p*=00.00,*C*.*I*=2.01-16.85),*also the odds for knowingly and intentionally did not Follow the government SOPs of Social Distancing During sharing of bedroom and toilet with positive symptomatic case* was 5.67 times more among the PCR Positive Contacts as compare to PCR negative Contacts(*p*=00.00,*C*.*I*=2.58-12.48), more over the odds for used the same vehicle after it was *used by the COVID-19 index case* was 19.58 times more among the PCR Positive Contacts as compare to PCR negative Contacts (*p*=00.00,*C*.*I*=10.19-37.60), also *the odds for Spoke with Positive COVID-19 index case for more than 15mins few days before catching the disease* was 15.58 times more among the PCR Positive Contacts as compare to the PCR negative Contacts (*p*=00.00,*C*.*I*=8.58-28.27), similarly the *odds for Individual did not use a face mask during all of his contact episodes with the positive index case* was 19.19 times more among the PCR Positive Contacts as compare to PCR negative Contacts(*p*=00.00,*C*.*I*=9.11-40.41), lastly the odds for *Participating in a gathering or social events* were 25.75 times more among the PCR Positive Contacts as compare to PCR negative Contacts(*p*=00.00,*C*.*I*=13.48-49.17).

In a similar study done by Hsiang S and the others^2^ where they have assessed the effectiveness of various ongoing anti-contagion policies, similar to our study results they have also reported positive effectiveness of various anti-contagious policies it was observed by them if SOPs and IPC measures were properly implemented it slowed the average growth rate of infections by − 0.252 per day (SE= 0.045, 164 p< 0.001) in China, − 0.248 (SE= 0.089, p< 0.01) in South Korea, − 0.24 (SE= 0.068, p< 0.001) in 165 Italy, − 0.355 (SE= 0.063, p< 0.001) in Iran, − 0.123 (SE= 0.019, p< 0.001) in France and − 0.084 166 (SE= 0.03, p< 0.01) in the US^2^.

Similarly a study by Lai S and others have predicted the infection rates and quantified the impact of various non-pharmacological interventions (NPI) among communities according their calculations Without NPIs, their model predicted the number of cases of COVID-19 to increase rapidly across China, with a 51-fold (IQR 33–71) increase in Wuhan, a 92-fold (58– 133) increase in other cities in Hubei province and a 125-fold (77–180) increase in other provinces by 29 February 2020. However, the apparent effectiveness of different interventions varied. Nevertheless, if intercity travel restrictions had been implemented, cities and provinces outside of Wuhan would have not received more cases from Wuhan, and the affected geographical range would not have expanded to the remote western areas of China. In general, they estimated that the early detection and isolation of cases quickly and substantially adopting IPC measure more infections were controlled like contact reduction and social distancing measures across the country. However, without the contact reduction intervention, in the longer term the epidemics would have increased exponentially across regions. Therefore, collective NPIs would bring about the strongest and most rapid effect on containment of the COVID-19 outbreak, with an interval of about one week between the introduction of NPIs and the peak of the epidemic^3^. The same effectiveness of Non pharmacological interventions like infection prevention and controls (IPC) measures if properly adopted the COVID-19 transmission rate could be lowered.

Similarly in another study by Flaxman S and the others showed that major non-pharmaceutical interventions (NPI)-and lockdowns in particular-have had a large effect on reducing transmission in Europeans. Continued intervention should be considered to keep transmission of SARS-CoV-2 under control^4^. Lockdown also causes social distancing similar to our study, social distancing is proved to be effective against COVID-19 transmission by both the studies.

Our this study is different from other such studies because we have studied the effectiveness of COVID-19 infection prevention and controls (IPC) measures among the contacts of an index case and have shown that the contacts who had positive COVID-19 PCR reports were poorly following the IPC measures.

## Conclusion & Recommendations

Being the first study of its kind in Pakistan the major findings of this study show that the PCR Positive contacts poorly adopted certain anti-COVID-19 IPC measures in their daily life hence got infected based on this evidence it is highly recommended that the media cell of the health department of the government of Balochistan should communicate the importance of these IPC measures to every individual of Balochistan and tell them their importance and adoption on regular bases in their daily life till the development and availability of COVID-19 vaccine for everyone.

## Data Availability

The data can be shared upon request.

## References

1 WHO. WHO coronavirus disease (COVID-19) dashboard. 2020. https://covid19.who.int (accessed Dec 15, 2020).

2 Hsiang S, Allen D, Annan-Phan S, et al. The effect of large-scale anti-contagion policies on the COVID-19 pandemic. Nature 2020; 584: 262–67.

3 Lai S, Ruktanonchai NW, Zhou L, et al. Effect of non-pharmaceutical interventions to contain COVID-19 in China. Nature 2020; 585: 410–13.

4 Flaxman S, Mishra S, Gandy A, et al. Estimating the effects of non-pharmaceutical interventions on COVID-19 in Europe. Nature 2020; 584: 257–61.

5 Arons MM, Hatfield KM, Reddy SC, et al. Presymptomatic SARS-CoV-2 infections and transmission in a skilled nursing facility. N Engl J Med 2020; 382: 2081–90.

7 Gudbjartsson DF, Helgason A, Jonsson H, et al. Spread of SARS-CoV-2 in the Icelandic population. N Engl J Med 2020; 382: 2302–15.

8 McMichael TM, Currie DW, Clark S, et al. Epidemiology of Covid-19 in a long-term care facility in King County, Washington. N Engl J Med 2020; 382: 2005–11.

9 Hamner L, Dubbel P, Capron I, et al. High SARS-CoV-2 attack rate following exposure at a choir practice — Skagit County, Washington, March 2020. MMWR Morb Mortal Wkly Rep 2020; 69: 606–10.

10 Luo L, Liu D, Liao X, et al. Contact settings and risk for transmission in 3410 close contacts of patients with COVID-19 in Guangzhou, China: a prospective cohort study. Ann Intern Med 2020; published online Aug 13. https://doi.org/10.7326/M20-2671.

11 Yong SEF, Anderson DE, Wei WE, et al. Connecting clusters of COVID-19: an epidemiological and serological investigation. Lancet Infect Dis 2020; 20: 809–15.

12 Sun Y, Koh V, Marimuthu K, et al. Epidemiological and clinical predictors of COVID-19. Clin Infect Dis 2020; 71: 786–92.

13 Ng Y, Li Z, Chua YX, et al. Evaluation of the effectiveness of surveillance and containment measures for the first 100 patients with COVID-19 in Singapore—January 2–February 29, 2020. MMWR Morb Mortal Wkly Rep 2020; 69: 307–11.

14 WHO. Coronavirus disease (COVID-19) advice for the public. 2020. https://www.who.int/emergencies/diseases/novel-coronavirus-2019/advice-for-public (accessed Sept 9, 2020).

15 Wei WE, Li Z, Chiew CJ, Yong SE, Toh MP, Lee VJ. Presymptomatic transmission of SARS-CoV-2—Singapore, January 23–March 16, 2020. MMWR Morb Mortal Wkly Rep 2020; 69: 411–15.

